# Artificial Intelligence Approximates Human Affect Ratings of Cannabis Images

**DOI:** 10.1101/2025.11.14.25339699

**Authors:** Jacob T. Borodovsky, Richard J. Macatee, Sarah M. Preum, Caroline L. Chung, Porter Malone, Denisse P. Gonzalez-Marquez

**Author notes:** Corresponding Author: Jacob T. Borodovsky, PhD., 46 Centerra Pkwy, Lebanon, NH 03766.

## Abstract

Cannabis imagery is proliferating online and can elicit affective responses related to use. Scalable tools are needed to evaluate how this proliferation could influence population health. This pilot study tested whether multimodal generative artificial intelligence (MGAI) can reproduce subjective human affect ratings of cannabis images. Four MGAI agents (model: gpt-4o-2024-11-20) were created to parallel the four human participant subgroups from Macatee et al. 2021, defined by primary method of cannabis administration (bong, bowl, joint/blunt, vaporizer). Using Macatee et al.’s participant instructions and standardized image set, each agent rated images of its primary method of administration on valence, arousal, and urge constructs. For each image-construct pair, n=100 ratings were generated in separate conversational threads using zero-shot prompting. Image-level MGAI mean ratings were compared with human mean ratings using Two One-Sided Tests of equivalence and Spearman correlations. Although formal statistical equivalence was rare (4% valence, 11% arousal, 3% urge), MGAI ratings approximated human ratings closely (Mean difference of mean ratings = – 0.31, SD = 1.23) and correlations between MGAI and human mean ratings were moderate to high: r_s_(valence) = 0.55, r_s_(arousal) = 0.34, r_s_(urge) = 0.56. MGAI also reproduced the parabolic relation between rating means and standard deviations observed in human data. These preliminary results indicate that MGAI can approximate human cannabis cue-reactivity patterns closely enough to justify continued refinement. MGAI could potentially be developed into a Cannabis Regulatory Science tool to aid regulatory oversight of online cannabis marketing.

## INTRODUCTION

The US legal cannabis industry continues to expand its e-commerce sales and online advertising. Consequently, images of cannabis-related products and paraphernalia (e.g., bud, vapes, edibles), brands, logos, and lifestyles have become pervasive online (Berg et al., 2023; Carlini et al., 2022; Cavazos-Rehg et al., 2019; Duan et al., 2023). When individuals use cannabis, the sensory stimuli present during use (e.g., sight of cannabis bud, lighter, pipe) become associated with the rewarding effects of intoxication and come to function as conditioned cues that independently elicit craving. When these cues are depicted in digital images, they can re-activate conditioned associations and elicit similar craving responses (Aston et al., 2015; Cousijn et al., 2013; Field et al., 2006; Field & Cox, 2008; Henry et al., 2014; Lundahl & Greenwald, 2015, 2016; McRae-Clark et al., 2011; Metrik et al., 2016; Norberg et al., 2016; Ruglass et al., 2019; Strickland et al., 2019; Vafaie & Kober, 2022; Wölfling et al., 2008). Marketers are aware of these dynamics and leverage them in advertising strategies to shape affective responses (Martin et al., 2013; Wang et al., 2023).

Compliance with cannabis advertising regulations remains low across many states with legal markets (Cao et al., 2020; Carlini et al., 2022; Marinello et al., 2024; Moreno et al., 2022; Sheikhan et al., 2021). The rapid expansion and decentralized structure of online cannabis marketing make oversight difficult and allow noncompliant content to persist (Caputi et al., 2018; Y. Cui et al., 2024; Duan et al., 2023). As a result, advertising often continues to feature salient cues that leverage use-related associations. Scalable tools that can navigate large digital ecosystems are therefore needed for effective monitoring of online cannabis imagery.

Multimodal generative artificial intelligence (MGAI) could be useful for regulatory oversight of aggressive online cannabis marketing. Recent studies show that MGAI can identify alcohol and nicotine products in digital images (Bonela et al., 2023; Kuntsche et al., 2023; Vassey et al., 2025). These studies represent important progress, but they have not focused specifically on cannabis imagery and have emphasized product detection rather than psychological impact. There is growing evidence that MGAI models can reasonably approximate various dimensions of human psychology (Argyle et al., 2023; Bail, 2024; Z. Cui et al., 2025). If MGAI can reproduce human affective responses to cannabis cues, it could potentially be used to help regulators triage review of online advertisements.

The purpose of the present study was to understand the extent to which, and ways in which, MGAI can approximate aggregate human affective ratings of cannabis cues. We instructed MGAI to provide ratings of valence, arousal, and urge for images in Macatee et al.’s (2021) standardized cannabis cue image set (Macatee et al., 2021). MGAI ratings were then compared with the corresponding human data reported in Macatee et al. We hypothesized that MGAI mean ratings would show statistical equivalence and strong rank order correlation with human mean ratings.

## METHODS

### Original Study Design

Macatee et al. (2021) developed and validated a standardized cannabis cue image set for cue-reactivity research. The image set contained 280 cannabis-related images and 80 neutral tooth-brushing images matched on visual and motor characteristics. Twenty cannabis images depicted cannabis flower alone, and 260 images depicted four methods of administration: bong, bowl, joint/blunt, and vaporizer. Each method-of-administration set consisted of 60 images distributed across three subtypes (20 per subtype): object-only images (e.g., bong shown alone), object-with-hand images, and object-with-face images (e.g., bong being used). The vaporizer category included 20 extra images to capture the diversity of vape products available. Participants were assigned to one of four subgroups based on their primary method of administration. Each participant rated the neutral and cannabis-flower images, and the images corresponding to their primary method (e.g., bong users rated bong images). Participants rated each image three times, once for each of the three constructs: valence, arousal, and urge. Ratings used a 1–9 Self-Assessment Manikin scale (Bradley & Lang, 1994) with labeled anchors at 1, 5, and 9. The present study used the object-only subset of method-of-administration images. MGAI agents provided ratings on the same constructs and scale, and MGAI mean ratings were compared with human mean ratings of the same object-only subset of images.

### MGAI Agents

We created four MGAI agents to parallel the four Macatee participant subgroups (bong, bowl, joint/blunt, vaporizer). Each agent had fixed “system instructions” that reflected the aggregate cannabis use characteristics (primary method of use, age at first use, age at regular use, and years of regular use; Figure 1, “Agent Setup”) of the corresponding human subsample from the Macatee study. Note that system instructions are different from the “prompt instructions” given to the agent. Verbatim system instructions for all four agents are provided in the appendix. All agents used GPT-4o (model: gpt-4o-2024-11-20), default hyperparameter values, and were queried via the OpenAI® “Assistants” API.

**Figure 1.**
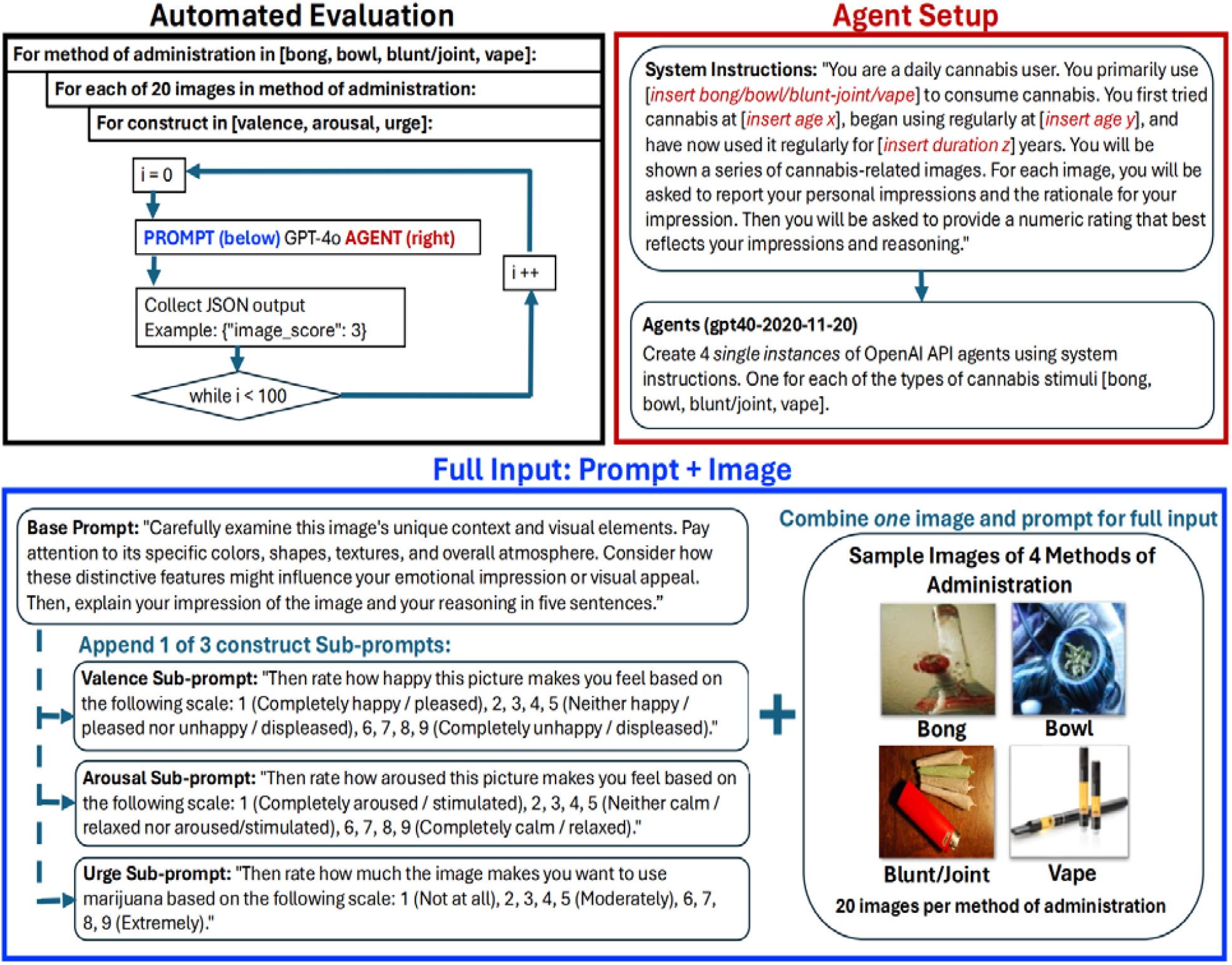
Study Procedures for Obtaining MGAI Ratings of Cannabis Images.

### Rating Procedure

We used the same three constructs (valence, arousal, urge) and the same 1–9 rating scale with labeled anchors as Macatee et al., but presented the scale to the agent as part of the prompt text. Each agent rated 20 images in its corresponding method-of-administration category, and the wording of the prompts mirrored the instructions given to Macatee et al. participants. An agent rated each image on each construct in 100 trials (i.e., n = 300 trials per image). For example, the Bong User agent rated bong image #1 in 100 trials on the valence scale, 100 trials on the arousal scale, and 100 trials on the urge scale, and then repeated this procedure for each of the other 19 bong images (Figure 1, “Automated Evaluation”). Each trial was executed as a unique “thread” to ensure that the agent had no memory of its ratings from prior trials. For each trial, the agent was provided with the cannabis image and prompt. The prompt contained three types of instructions: (i) inspect image-specific features, (ii) generate a short rationale, and (iii) use a construct-specific rating question with 1–9 anchors to rate the image. Full prompt templates are provided in Figure 1 (“Full Input, Prompt + Image”) and the appendix.

### Statistical Analysis

We first tested statistical equivalence of MGAI and human mean ratings at the image–construct level using Two One-Sided Tests (TOST). For each image–construct pair, we calculated the pooled standard deviation of MGAI trial-level ratings and human participant-level ratings. We multiplied the pooled standard deviation by 0.5 to define equivalence bounds (k = 0.5). We set α = 0.05. Equivalence required both one-sided p-values < 0.05 and a 90% confidence interval that fell entirely within the equivalence bounds. We planned 240 equivalence tests (20 images × 3 constructs × 4 method-of-administration categories). For example, the mean MGAI valence rating for bong image #1 was compared to the mean human valence rating for bong image #1. One test (valence for images V7) could not be conducted because MGAI trial-level standard deviations equaled zero. No alpha corrections were applied as we treated each image-construct comparison as an individual hypothesis (Rubin, 2021). We also compared MGAI and human ratings using Spearman correlation (r_s_) within each construct, and within each method-of-administration category. All analyses were performed in R using dplyr for data wrangling, purrr for iteration, and TOSTER for equivalence testing. This study was deemed exempt from human subjects review by the Dartmouth Committee for the Protection of Human Subjects.

## RESULTS

TOST-based equivalence of Human and MGAI ratings was rare. Overall, 4%, 11%, and 3% of MGAI’s mean valence, arousal, and urge ratings, respectively, were statistically equivalent to human mean ratings. When stratified by method of administration, approximately 5%, 8%, 3%, and 7% of MGAI’s mean ratings for bowl, vape, blunt/joint, and bong, respectively, were statistically equivalent to human mean ratings. We also observed that approximately 98%, 90%, and 3% of the differences (MGAI mean minus human mean) in valence, arousal, and urge ratings, respectively, were negative, indicating that MGAI consistently underestimated valence and arousal ratings and overestimated urge ratings.

Despite the lack of TOST-based equivalence, differences between MGAI mean ratings and human mean ratings were small (mean difference of means = -0.308, SD = 1.230)(Figure 2a), and the Spearman correlations between MGAI mean ratings and human mean ratings were high: valence (r_s_ = 0.552), arousal (r_s_ = 0.335), and urge (r_s_ = 0.562). When stratified by method of administration, Spearman correlations between MGAI mean ratings and human mean ratings were: bowl (r_s_ = 0.203), vape (r_s_ = 0.004), blunt/joint (r_s_ = 0.287), and bong (r_s_ = 0.441). Notably, MGAI rating standard deviations were markedly lower than human rating standard deviations. Average standard deviations were: valence (MGAI: 0.340 vs. human: 2.019), arousal (MGAI: 1.164 vs. human: 2.428), and urge (MGAI: 0.561 vs. human: 2.341). However, the relationship between the mean and standard deviation of ratings showed similar parabolic relationships for MGAI and humans (Figure 2b).

**Figure 2.**
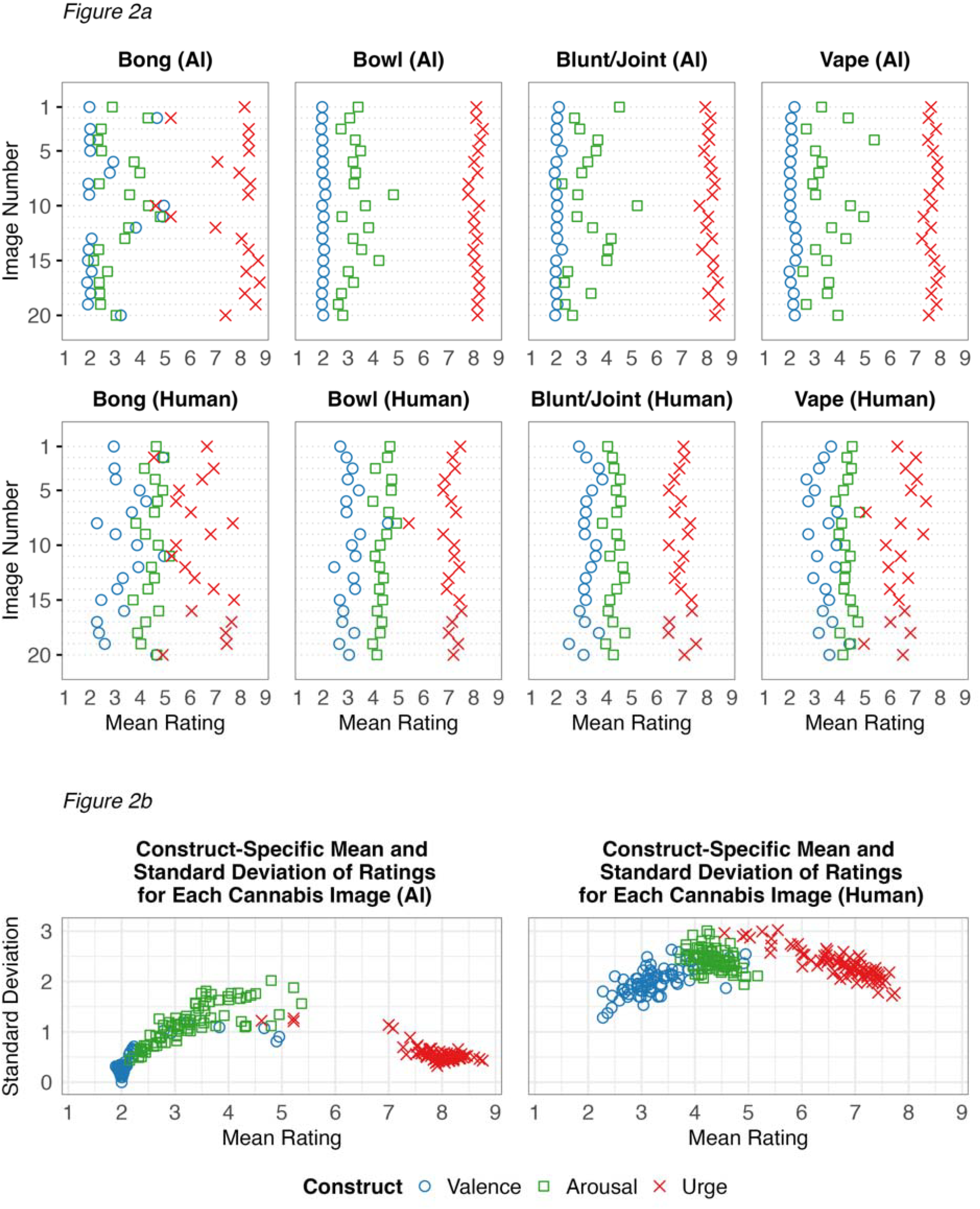
(2A) Mean Ratings per Image by Source and Product Category; (2B) Scatterplot of Mean and Standard Deviation of Ratings by Source.

## DISCUSSION

We tested whether multimodal generative artificial intelligence (MGAI) could reproduce human ratings of cannabis images across four methods of administration (bong, bowl, joint/blunt, vaporizer) on three psychological constructs (valence, arousal, urge). Statistical equivalence of mean MGAI ratings and mean human ratings was rare. However, MGAI–human differences in mean ratings were small, and the rank ordering of mean ratings of constructs observed in humans (valence < arousal < urge) was well replicated by MGAI both overall and within method-of-administration categories. Thus, the lack of statistical equivalence seems to be due to smaller standard deviations of MGAI ratings rather than large MGAI–human discrepancies in mean ratings. Although further work is needed to clarify the sources of MGAI–human discrepancies, these findings provide initial evidence that MGAI could be developed into a Cannabis Regulatory Science (Borodovsky et al., 2021; Borodovsky & Budney, 2018) tool for assisting regulatory review of online cannabis advertising.

Across method-of-administration categories, MGAI preserved the relative rank order pattern of mean ratings of constructs observed in the human data. However, MGAI systematically “amplified” that pattern by producing mean ratings farther from the midpoint of the scale (Figure 2a). Several recent studies have reported similar findings (Alrasheed et al., 2025; Romeo & Testolin, 2025; Shirahama et al., 2025). Despite this amplification bias, MGAI still reproduced the parabolic relation between rating means and standard deviations observed in our human sample and other human samples (Pollock, 2018). This correspondence is noteworthy because, in principle, MGAI could have produced mean ratings at the scale midpoint with narrow standard deviations (e.g., MGAI rates the image as a 5 in all 100 trials).

Several limitations qualify these findings. First, procedural differences between the MGAI and human tasks may have influenced results. MGAI agents were required to produce a five-sentence rationale before each rating (Chu et al., 2024) and used text-based anchors, whereas participants in Macatee et al. (2021) were not asked for a rationale and provided ratings using Self-Assessment Manikin pictorial scales. Second, all 100 MGAI trials for a given image– construct pairing originated from the same agent identity across separate threads. Using a larger pool of independent agents, with each agent providing a single rating per image, could have better approximated the distributions of independent human raters. Third, in the absence of a field standard, we relied on a conservative k = 0.5 bound for our TOST analysis, which strongly influences whether a result is labeled “equivalent”. Fourth, we evaluated only a single MGAI platform and model (ChatGPT, version GPT-4o-2024-11-20); performance may differ across other commercially available MGAI systems (e.g., Gemini, Claude) or future OpenAi ® models.

These preliminary findings indicate that MGAI can approximate average human cannabis cue-reactivity patterns closely enough to justify continued methodological testing and refinement. With further development under a Cannabis Regulatory Science framework, these tools could be used to aid Federal or State oversight of online cannabis-based product advertisements (Food and Drug Administration, 2025; Padon et al., 2025) that violate public health regulations.

## Data Availability

All data produced in the present study are available upon reasonable request to the authors

## Acknowledgments

Funding for this study was provided by grants from the National Institute on Drug Abuse (NIDA) R01DA050032, R21DA062816, P30DA037202. The funding organizations had no role in the design and conduct of the study; collection, management, analysis, and interpretation of the data; preparation, review, or approval of the manuscript; and decision to submit the manuscript for publication. Author JTB employed ChatGPT and Grammarly to help refine grammar, sentence structure, and word choice. The author(s) reviewed and retain full responsibility for the manuscript’s content.

## BIBLIOGRAPHY

Alrasheed, H., Alghihab, A., Pentland, A., & Alghowinem, S. (2025). Evaluating the capacity of large language models to interpret emotions in images. PLOS ONE, 20(6), e0324127. 10.1371/journal.pone.0324127

Argyle, L. P., Busby, E. C., Fulda, N., Gubler, J. R., Rytting, C., & Wingate, D. (2023). Out of One, Many: Using Language Models to Simulate Human Samples. Political Analysis, 31(3), 337–351. 10.1017/pan.2023.2

Aston, E. R., Metrik, J., & MacKillop, J. (2015). Further validation of a marijuana purchase task. Drug and Alcohol Dependence, 152, 32–38. 10.1016/j.drugalcdep.2015.04.025

Bail, C. A. (2024). Can Generative AI improve social science? Proceedings of the National Academy of Sciences, 121(21), e2314021121. 10.1073/pnas.2314021121

Berg, C. J., Romm, K. F., Pannell, A., Sridharan, P., Sapra, T., Rajamahanty, A., Cui, Y., Wang, Y., Yang, Y. T., & Cavazos-Rehg, P. A. (2023). Cannabis retailer marketing strategies and regulatory compliance: A surveillance study of retailers in 5 US cities. Addictive Behaviors, 143, 107696. 10.1016/j.addbeh.2023.107696

Bonela, A. A., Nibali, A., He, Z., Riordan, B., Anderson-Luxford, D., & Kuntsche, E. (2023). The promise of zero-shot learning for alcohol image detection: Comparison with a task-specific deep learning algorithm. Scientific Reports, 13(1), 11891. 10.1038/s41598-023-39169-4

Borodovsky, J. T., & Budney, A. J. (2018). Cannabis regulatory science: Risk-benefit considerations for mental disorders. Int Rev Psychiatry, 30(3), 183–202. 10.1080/09540261.2018.1454406

Borodovsky, J. T., Sofis, M. J., Grucza, R. A., & Budney, A. J. (2021). The importance of psychology for shaping legal cannabis regulation. Exp Clin Psychopharmacol, 29(1), 99– 115. 10.1037/pha0000362

Bradley, M. M., & Lang, P. J. (1994). Measuring emotion: The self-assessment manikin and the semantic differential. Journal of Behavior Therapy and Experimental Psychiatry, 25(1), 49–59. 10.1016/0005-7916(94)90063-9

Cao, Y., Carrillo, A. S., Zhu, S., & Shi, Y. (2020). Point-of-Sale Marketing in Recreational Marijuana Dispensaries Around California Schools. Journal of Adolescent Health, 66(1), 72–78. 10.1016/j.jadohealth.2019.07.023

Caputi, T. L., Leas, E. C., Dredze, M., & Ayers, J. W. (2018). Online Sales of Marijuana: An Unrecognized Public Health Dilemma. American Journal of Preventive Medicine, 54(5), 719–721. 10.1016/j.amepre.2018.01.032

Carlini, B. H., Garrett, S., Firth, C., & Pinsky, I. (2022). Cannabis Industry Marketing Violations in Washington State, 2014-2019. Journal of Studies on Alcohol and Drugs, 83(1), 18–26. 10.15288/jsad.2022.83.18

Cavazos-Rehg, P. A., Krauss, M. J., Cahn, E., Lee, K. E., Ferguson, E., Rajbhandari, B., Sowles, S. J., Floyd, G. M., Berg, C., & Bierut, L. J. (2019). Marijuana Promotion Online: An Investigation of Dispensary Practices. Prevention Sciencel: The Official Journal of the Society for Prevention Research, 20(2), 280–290. 10.1007/s11121-018-0889-2

Chu, K., Chen, Y.-P., & Nakayama, H. (2024). A Better LLM Evaluator for Text Generation: The Impact of Prompt Output Sequencing and Optimization. 10.11517/pjsai.JSAI2024.0_2G5GS604

Cousijn, J., Goudriaan, A. E., Ridderinkhof, K. R., van den Brink, W., Veltman, D. J., & Wiers, R. W. (2013). Neural responses associated with cue-reactivity in frequent cannabis users. Addict Biol, 18(3), 570–580. 10.1111/j.1369-1600.2011.00417.x

Cui, Y., Duan, Z., LoParco, C. R., Vinson, K., Romm, K. F., Wang, Y., Cavazos-Rehg, P. A., Kasson, E., Yang, Y. T., & Berg, C. J. (2024). Changes in online marketing and sales practices among non-medical cannabis retailers in 5 US cities, 2022 to 2023. Preventive Medicine Reports, 42, 102755. 10.1016/j.pmedr.2024.102755

Cui, Z., Li, N., & Zhou, H. (2025). A large-scale replication of scenario-based experiments in psychology and management using large language models. Nature Computational Science, 1–8. 10.1038/s43588-025-00840-7

Duan, Z., Kasson, E., Ruchelli, S., Rajamahanty, A., Williams, R., Sridharan, P., Sapra, T., Dopke, C., Pannell, A., Nakshatri, S., Berg, C. J., & Cavazos-Rehg, P. A. (2023). Assessment of Online Marketing and Sales Practices Among Recreational Cannabis Retailers in Five U.S. Cities. Cannabis and Cannabinoid Research. 10.1089/can.2022.0334

Field, M., & Cox, W. M. (2008). Attentional bias in addictive behaviors: A review of its development, causes, and consequences. Drug Alcohol Depend, 97(1–2), 1–20. 10.1016/j.drugalcdep.2008.03.030

Field, M., Eastwood, B., Bradley, B. P., & Mogg, K. (2006). Selective processing of cannabis cues in regular cannabis users. Drug Alcohol Depend, 85(1), 75–82. 10.1016/j.drugalcdep.2006.03.018

Food and Drug Administration. (2025, September 5). Warning Letters for Cannabis-Derived Products. FDA. https://www.fda.gov/news-events/public-health-focus/warning-letters-cannabis-derived-products

Henry, E. A., Kaye, J. T., Bryan, A. D., Hutchison, K. E., & Ito, T. A. (2014). Cannabis cue reactivity and craving among never, infrequent and heavy cannabis users. Neuropsychopharmacology, 39(5), 1214–1221. 10.1038/npp.2013.324

Kuntsche, E., He, Z., Bonela, A. A., & Riordan, B. (2023). Zero-shot learning has the potential to revolutionise research on exposure to alcohol and other drugs in digital media. International Journal of Drug Policy, 118, 104098. 10.1016/j.drugpo.2023.104098

Lundahl, L. H., & Greenwald, M. K. (2015). Effect of oral THC pretreatment on marijuana cue-induced responses in cannabis dependent volunteers. Drug Alcohol Depend, 149, 187– 193. 10.1016/j.drugalcdep.2015.01.046

Lundahl, L. H., & Greenwald, M. K. (2016). Magnitude and duration of cue-induced craving for marijuana in volunteers with cannabis use disorder. Drug Alcohol Depend, 166, 143– 149. PubMed. 10.1016/j.drugalcdep.2016.07.004

Macatee, R. J., Carr, M., Afshar, K., & Preston, T. J. (2021). Development and validation of a cannabis cue stimulus set. Addictive Behaviors, 112, 106643. 10.1016/j.addbeh.2020.106643

Marinello, S., Valek, R., & Powell, L. M. (2024). Analysis of social media compliance with cannabis advertising regulations: Evidence from recreational dispensaries in Illinois 1-year post-legalization. Journal of Cannabis Research, 6(1), 2. 10.1186/s42238-023-00208-6

Martin, I. M., Kamins, M. A., Pirouz, D. M., Davis, S. W., Haws, K. L., Mirabito, A. M., Mukherjee, S., Rapp, J. M., & Grover, A. (2013). On the road to addiction: The facilitative and preventive roles of marketing cues. Journal of Business Research, 66(8), 1219– 1226. 10.1016/j.jbusres.2012.08.015

McRae-Clark, A. L., Carter, R. E., Price, K. L., Baker, N. L., Thomas, S., Saladin, M. E., Giarla, K., Nicholas, K., & Brady, K. T. (2011). Stress- and cue-elicited craving and reactivity in marijuana-dependent individuals. Psychopharmacology (Berl), 218(1), 49–58. 10.1007/s00213-011-2376-3

Metrik, J., Aston, E. R., Kahler, C. W., Rohsenow, D. J., McGeary, J. E., Knopik, V. S., & MacKillop, J. (2016). Cue-elicited increases in incentive salience for marijuana: Craving, demand, and attentional bias. Drug and Alcohol Dependence, 167, 82–88. 10.1016/j.drugalcdep.2016.07.027

Moreno, M. A., Jenkins, M., Binger, K., Kelly, L., Trangenstein, P. J., Whitehill, J. M., & Jernigan, D. H. (2022). A Content Analysis of Cannabis Company Adherence to Marketing Requirements in Four States. Journal of Studies on Alcohol and Drugs, 83(1), 27–36. 10.15288/jsad.2022.83.27

Norberg, M. M., Kavanagh, D. J., Olivier, J., & Lyras, S. (2016). Craving cannabis: A meta-analysis of self-report and psychophysiological cue—reactivity studies. Addiction, 111(11), 1923–1934. 10.1111/add.13472

Padon, A. A., Ghahremani, D. G., Simard, B., Soroosh, A. J., & Silver, L. D. (2025). Characteristics and effects of cannabis advertisements with appeal to youth in California. International Journal of Drug Policy, 137, 104718. 10.1016/j.drugpo.2025.104718

Pollock, L. (2018). Statistical and methodological problems with concreteness and other semantic variables: A list memory experiment case study. Behavior Research Methods, 50(3), 1198–1216. 10.3758/s13428-017-0938-y

Romeo, Z., & Testolin, A. (2025). Artificial intelligence can emulate human normative judgments on emotional visual scenes. Royal Society Open Science, 12(7), 250128. 10.1098/rsos.250128

Rubin, M. (2021). When to adjust alpha during multiple testing: A consideration of disjunction, conjunction, and individual testing. Synthese, 199(3), 10969–11000. 10.1007/s11229-021-03276-4

Ruglass, L. M., Shevorykin, A., Dambreville, N., & Melara, R. D. (2019). Neural and behavioral correlates of attentional bias to cannabis cues among adults with cannabis use disorders. Psychol Addict Behav, 33(1), 69–80. 10.1037/adb0000423

Sheikhan, N. Y., Pinto, A. M., Nowak, D. A., Abolhassani, F., Lefebvre, P., Duh, M. S., & Witek, T. J., Jr. (2021). Compliance With Cannabis Act Regulations Regarding Online Promotion Among Canadian Commercial Cannabis-Licensed Firms. JAMA Network Open, 4(7), e2116551. 10.1001/jamanetworkopen.2021.16551

Shirahama, N., Nakaya, N., Watanabe, S., Moriya, K., Matsumoto, K., & Koshi, K. (2025). Assessing Emotional Intelligence in AI: Journal of the Institute of Industrial Applications Engineers, 13(1), 21–21. 10.12792/jiiae.13.21

Strickland, J. C., Lile, J. A., & Stoops, W. W. (2019). Contribution of cannabis-related cues to concurrent reinforcer choice in humans. Drug and Alcohol Dependence, 199, 85–91. 10.1016/j.drugalcdep.2019.02.022

Vafaie, N., & Kober, H. (2022). Association of Drug Cues and Craving With Drug Use and Relapse: A Systematic Review and Meta-analysis. JAMA Psychiatry, 79(7), 641–650. 10.1001/jamapsychiatry.2022.1240

Vassey, J., Kennedy, C. J., Chang, H.-C. H., & Unger, J. B. (2025). Generative AI in a new era of computer model-informed tobacco research: A short report. Tobacco Control. 10.1136/tc-2024-058887

Wang, E. S.-T., Lin, J.-Y., & Liao, Y.-T. (2023). The Effects of Positive Valence and Intensity of Word-Of-Mouth and Advertising on Forming Beer Brand Equity and Purchase Intentions. Journal of Food Products Marketing, 29(8–9), 255–269. 10.1080/10454446.2023.2273517

Wölfling, K., Flor, H., & Grüsser, S. M. (2008). Psychophysiological responses to drug-associated stimuli in chronic heavy cannabis use. European Journal of Neuroscience, 27(4), 976–983. 10.1111/j.1460-9568.2008.06051.x

